# Using mobile phone data for epidemiological simulations of lockdowns: government interventions, behavioral changes, and resulting changes of reinfections

**DOI:** 10.1101/2020.07.22.20160093

**Authors:** Sebastian A Müller, Michael Balmer, Billy Charlton, Ricardo Ewert, Andreas Neumann, Christian Rakow, Tilmann Schlenther, Kai Nagel

## Abstract

Epidemiological simulations as a method are used to better understand and predict the spreading of infectious diseases, for example of COVID-19. This paper presents an approach that combines person-centric data-driven human mobility modelling with a mechanistic infection model and a person-centric disease progression model. Results show that in Berlin (Germany), behavioral changes of the population mostly happened *before* the government-initiated so-called contact ban came into effect. Also, the model is used to determine differentiated changes to the reinfection rate for different interventions such as reductions in activity participation, the wearing of masks, or contact tracing followed by quarantine-at-home. One important result is that successful contact tracing reduces the reinfection rate by about 30 to 40%, and that if contact tracing becomes overwhelmed then infection rates immediately jump up accordingly, making rather strong lockdown measures necessary to bring the reinfection rate back to below one.

## 1 Background

The general dynamics of virus spreading is captured by compartmental models, most famously the so-called SIR model, with *S* = *susceptible, I* = *infected/infectious*, and *R* = *recovered* [1, 2]. Every time a *susceptible* and an *infectious* person meet, there is a probability that the susceptible person becomes infected. Some time after the infection, the person typically recovers. Variants include, e.g., an *exposed* (but not yet *infectious*) compartment between *S* and *I*.

Compartmental models assume transition rates from one compartment to the other, most importantly the infection rate *β* for the transition from *S* to *I*. Smieszek [3, 4] presents a more realistic mechanical infection model: Infected persons generate a “viral load” that they exhale, cough or sneeze into the environment, and people close by are exposed. Overall, the probability for person *n* to become infected by this process in a time step *t* is described as

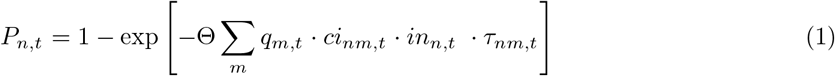

where *m* is a sum over all other persons, *q* is the shedding rate (~ microbial load), *ci* the contact intensity, *in* the intake (reduced, e.g., by a mask), and *τ* the duration of interaction between the two individuals.

Such models need to be embedded into a contact graph [5, 6, 7, 8]. An infected person from out-of-country may return to its family, there infect one of her/his children, the child may take it to the school where it infects other children, etc. These trajectories are typically not available for simulations, for example for privacy reasons. It is, however, possible to generate synthetic approximations to these trajectories. This is routinely done in transport modelling. One approach is to use information from mobile phone data (but not the full trajectories), and process them together with information about the transport system and with statistical information from other surveys [9, 10]. That approach leads to synthetic movement trajectories for the complete population (cf. Fig. 1). From these trajectories, it is possible to extract how much time people spend with other people at activities or in (public transport) vehicles.

**Figure 1.**
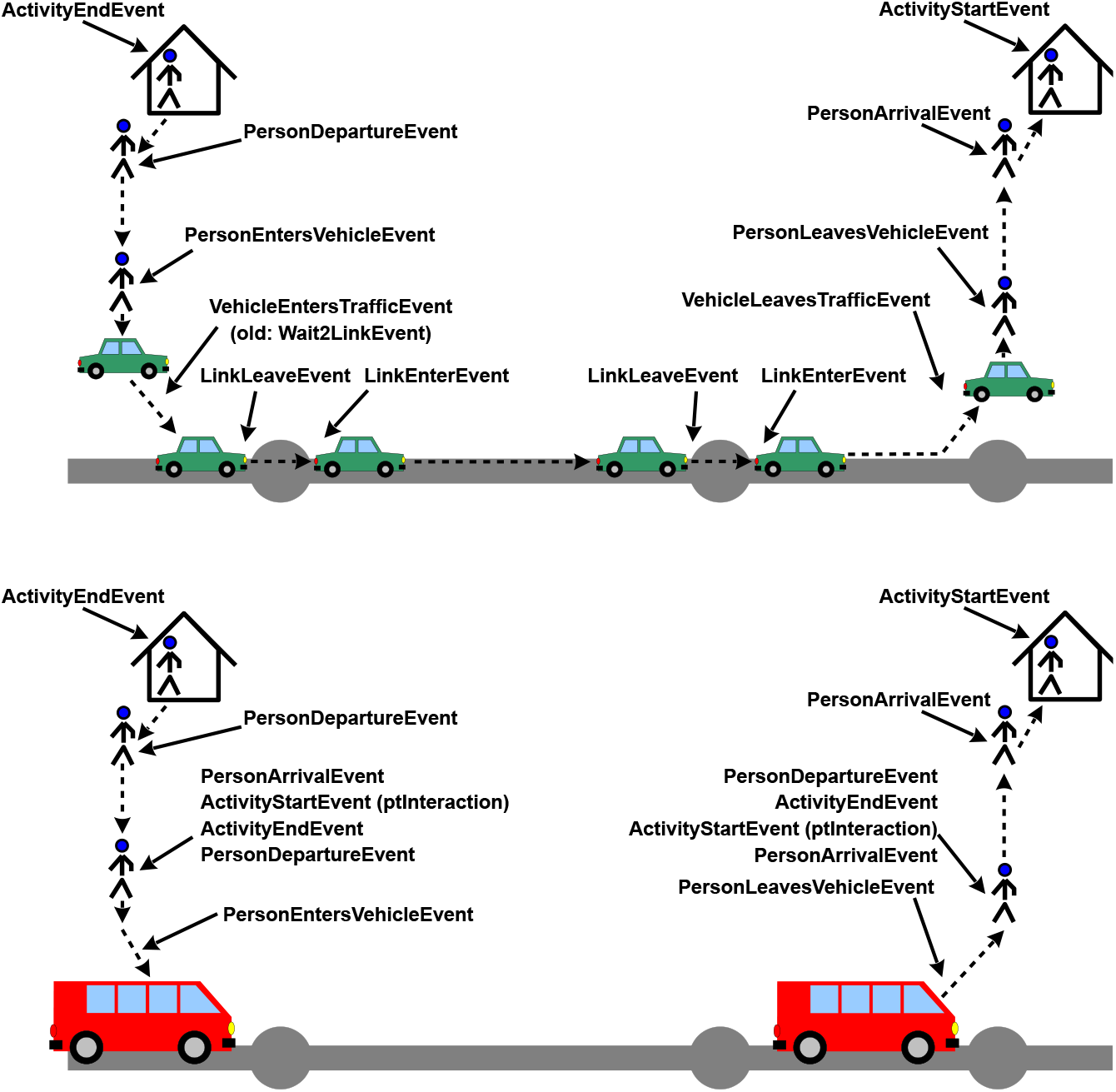
Top: Events for travel by individual vehicle. Bottom: Events for travel by public transport. Source: [11].

This paper presents simulation results for such a data set, for the metropolitan area of Berlin in Germany, with approx. 5 million people. The simulation model constructs an infection dynamics model on top of persons’ movement trajectories:

1. One or more *exposed* persons are introduced into the population.
2. At some point, *exposed* persons become *infectious*. From then on, every time they spend time together with some other person in a vehicle or at some activity, Eq. (1) is used to calculate the probability that the other person, if *susceptible*, can become infected (= *exposed*). If infection happens, the newly infected person will follow the same dynamics.
3. *Infectious* persons eventually move on to other states, described later.

The model runs many days, until no more infections occur.

This continues work by Smieszek et al [3, 12] and by Hackl and Dubernet [13]. The important difference, and major innovation, is that our model is entirely data-driven on the mobility side, i.e. both the “normal” person trajectories and the reduction of activity participation over the course of the epidemics stem from data. This allows to considerably reduce the number of free parameters.

Other similar work, i.e. virus spreading based on a mobility model, is by Virginia Biotechnology Institute [14, 15]. However, again, our model is much more data driven. The work by Imperial College [15, 16] is similar to ours in that it models individual persons. However, again, our model is stronger on the mobility side, and thus able to obtain compliance rates from data rather than having to guess them.

## 2 Methods

### 2.1 Disease progression model

The disease progression model is taken from the literature [17, 18, 19, 20, 21, 22] (also see [23]). The model has states *exposed, infectious, showing symptoms, seriously sick*(= should be in hospital), *critical* (= needs intensive care), and *recovered*. The durations from one state to the next follow log-normal distributions; see Fig. 2 for details. We use the same age-dependent transition probabilities as [16], shown in Tab. 1.

**Figure 2.**
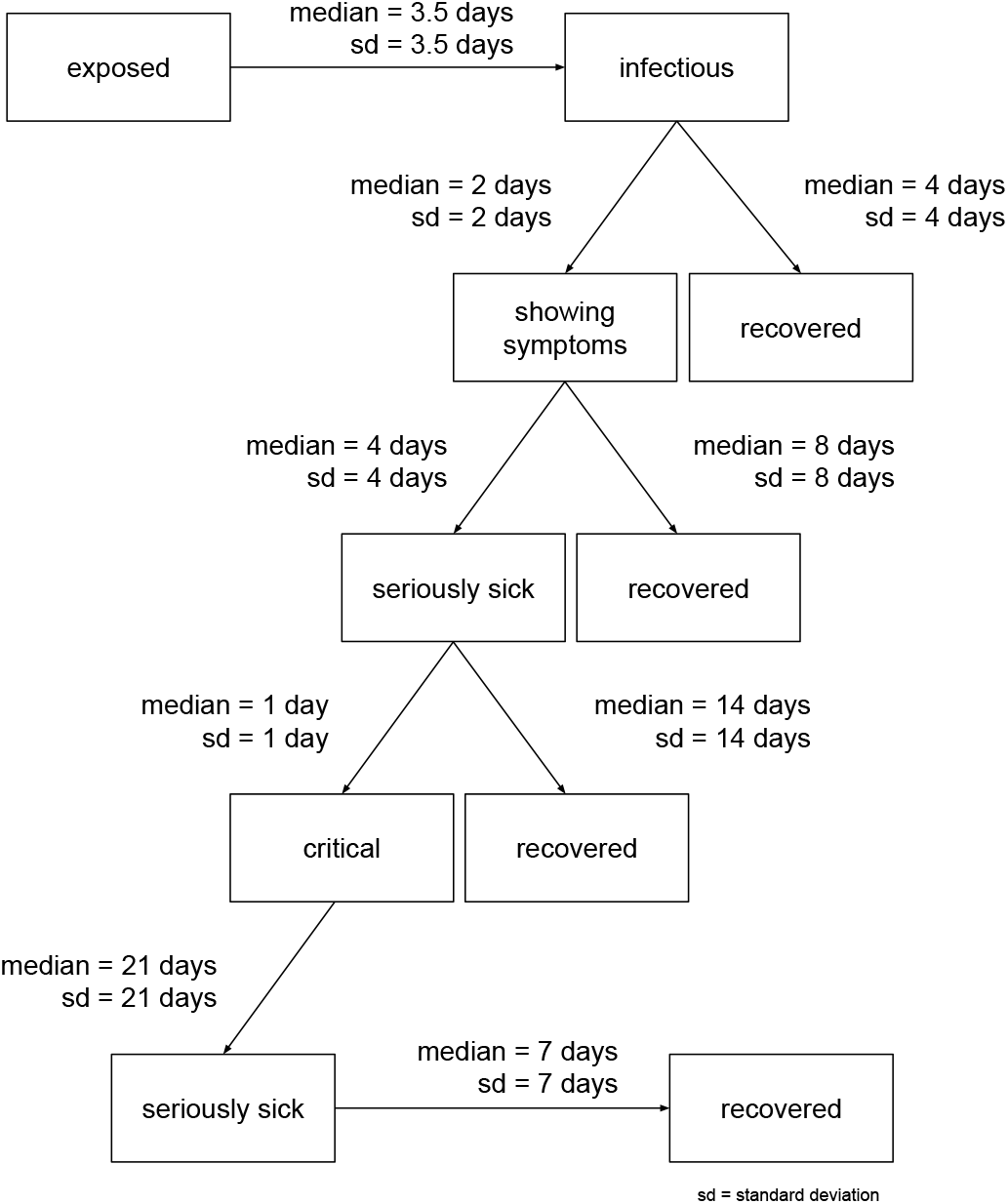
State transitions [17, 18, 19, 20, 21, 22],

**Table 1.**
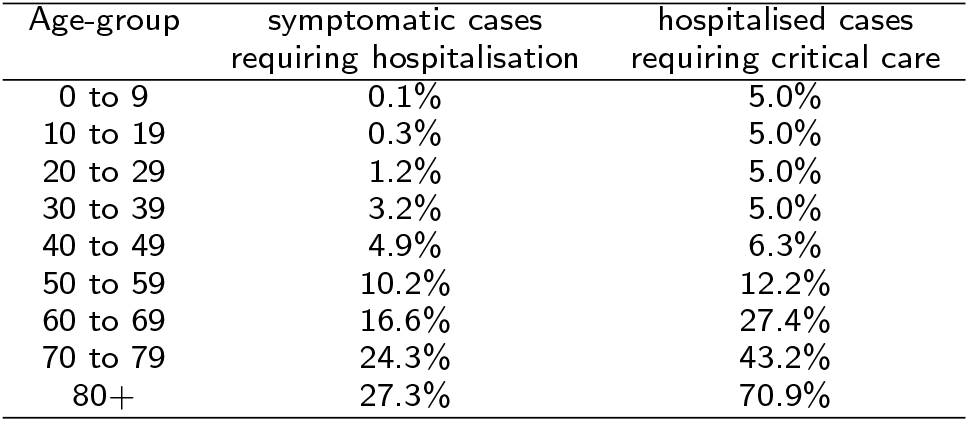
Age-dependent transition probabilities from symptomatic to seriously sick (= requiring hospitalisation), and from seriously sick to critical (= requiring breathing support or intensive care). Source: [16].

| Age-group | symptomatic cases<br>requiring hospitalisation | hospitalised cases<br>requiring critical care |
| --- | --- | --- |
| 0 to 9 | 0.1% | 5.0% |
| 10 to 19 | 0.3% | 5.0% |
| 20 to 29 | 1.2% | 5.0% |
| 30 to 39 | 3.2% | 5.0% |
| 40 to 49 | 4.9% | 6.3% |
| 50 to 59 | 10.2% | 12.2% |
| 60 to 69 | 16.6% | 27.4% |
| 70 to 79 | 24.3% | 43.2% |
| 80+ | 27.3% | 70.9% |

Infecting another person is possible during *infectious*, and while *showing symptoms*, but no longer than 4 days after becoming infectious. This models that persons are mostly infectious relatively early through the disease [18], while in later stages the infection may move to the lung [19], which makes it worse for the infected person, but seems to make it less infectious to other persons.

### 2.2 Contact model

Encounters in “containers”, i.e. in facilities and vehicles, are directly taken from the data. Synthetic persons in the same container at the same time have a certain probability to interact; if they interact, the probability of an infection is given by Eq. (1) if one person is contagious (maximally until 4 days after becoming *infectious*; see above) and the other is *susceptible*. In particular, the duration of the interaction to feed into Eq. (1) is also provided by the data.

The algorithm looks at agents when they *leave* a facility. At this point, it randomly selects 3 other agents which are at the facility at the same time, and with each selected agent computes a possible infection if either the leaving agent is susceptible and the other agent is contagious, or the other way round. The infection model is Eq. (1), the time τ is the time (duration) that both agents were simultaneously at the same facility. We bound the infection dynamics at 3 other agents, since we assume that persons do not interact with everybody in the facility. If there are fewer than 3 other agents at the facility, then interaction simply happens with everybody. Clearly, we do not want the model to interact with everybody at the facility. Using “10” instead of “3” in trial runs did not make a noticeable difference (except that the 0 parameter needed recalibration).

The same algorithm is used for interaction in (public transport) vehicles. In terms of implementation, the model uses a generalized dynamics for *containers*, and treats both facilities and vehicles as such containers.

Note that the number “3” is used when the agent leaves. There is, however, also interaction when other agents leave. Thus, this rather models the interaction with “available spaces” than with persons. E.g. assume the following time line (from left to right) and consider in particular the agent X:

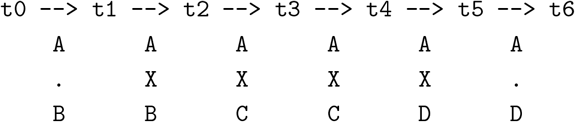

This can, for the ease of interpretation, be interpreted as three seats in a row in a public transport vehicle. We have the following events with respect to person X:

- At time 11, X enters the vehicle, and takes a seat between A and B.
- At time t2, B leaves the vehicle. In consequence, a possible infection of X from B is computed with Eq. (1), with *t*2 – *t*1 as the duration of the interaction. Also, C enters the vehicle, and takes the seat of B.
- At time *t*4, C leaves the vehicle. A possible infection of X from C is computed, with *t*4 –*t*2 as the interaction time. At the same time, D enters the vehicle, and takes the seat of C.
- At time *t*5, X leaves the vehicle. A possible infection of X from A is computed, with *t*5 –*t*1 as interaction time, and from D, with *t*5 –*t*4 as interaction time.

In the algorithm, however, we do not explicitly model seating positions, but just assume that every synthetic person that leaves interacts with up to 3 randomly selected other agents.

The variation over time of the density of persons inside the container is currently not taken into account. This could, however, be done in a further modelling step, given data of facility and vehicle sizes. What *is*, however, taken into account is the thinning out of persons in the container when they are reducing their activity participation (see Sec. 2.5) no longer there: The algorithm still computes interaction with up to 3 randomly selected persons, but if these persons do not participate at that activity on that day, then there is no infection dynamics in either direction.

### 2.3 Multi-day modelling

Optimally, one would have multi-day trajectories. In our case, the data that we have ends at the end of the day. Our simulations thus run the same person trajectories again and again. This presumably *underestimates* mixing. However, there is still strong mixing because the synthetic persons interact with other persons at the same facility every day. For example, a train may have 1000 passengers. Out of these, any of our agents would only see a small number other agents. This leads to different encounters in every synthetic day, even when repeating the same trajectory over and over.

The same holds for facilities, where the data is actually constructed such that there are up to 400 persons per facility. That is, any work, shopping, or leisure facility can have up to 400 visitors at the same time, and every person that has that facility in its trajectory can interact with all of these persons. This number of interacting agents on each single day is, however, limited (see infections at facilities and in vehicles). This results in an issue for home locations: Our data model does not differentiate between “persons living in the same block” and “persons living in the same household”. For this reason we split persons living in the same block into realistic household sizes with a maximum number of six people per household [24].

As stated before, activities are of a certain type. These types are used in this research to determine the effects that certain strategies have on the epidemic spreading. If an activity is removed from a schedule, agents will still be put in the respective containers, but do not take part in the infections dynamics. Such agents essentially represent “holes”, since they may still be picked as one of the three agents to interact with, but there is no infection.

### 2.4 Base calibration

Most parameters of the model are taken from the literature, as explained above and in the additional material. The remaining free parameters are, from Eq. (1), Θ · *q* · *ci* · *in*. Since none of these numbers seem to be known for COVID-19, we have first set the base values of *q* = *ci* = *in* = 1. We keep these parameters separate, since they later allow the modelling of different contact intensities and of masks. The remaining Θ parameter was then calibrated so that in the base case, the number of cases doubles every three days. This is a plausible fit to the initial days which were totally without distancing measures, which is what our base case represents. Calibrating Θ in our model corresponds to calibrating *β*, the infection rate, in a compartmental model.

Another free parameter is when the first infection occurs. This corresponds to the initial number of infected persons, *I_o_*, in the compartmental models. This date was set such that our number of persons showing symptoms, which corresponded to the sampling strategy in Germany during the early phase, is 50% above the reported case numbers. Evidently, this has to do with the fraction of non-reported cases, which is considered in Section 2.7. We also feed the simulation one additional infected case per day, since Berlin is embedded into a system where disease import is probable. The result of this base calibration can be seen in Fig. 5 (blue).

### 2.5 Reductions in activity participation

During the unfolding of the epidemics, people decided to no longer participate in certain activities. We model this by removing an activity from a person’s schedule, plus the travel to and from the activity. In consequence, that person no longer interacts with people at that activity location, and in consequence neither can infect other persons nor can become infected during that activity. Overall, this reduces contact options, and thus reduces epidemic spread.

A very important consequence of our modelling approach is that we can take that reduction in activity participation from data. Unfortunately, the activity type detection algorithm is not very good for these unusual activity patterns, as one can see in Fig. 3 when knowing that all educational institutions were closed in Berlin after Mar/15. What is reliable, though, is the differentiation between at-home and out-of-home time, as displayed in Fig. 4. One clearly notices that out-of-home activities are somewhat reduced after Mar/8, and dramatically reduced soon after. After some experimentation, it was decided to take weekly averages of the activity non-participation, and use that uniformly across all activity types in our model, except for educational activities, which were taken as ordered by the government.

**Figure 3.**
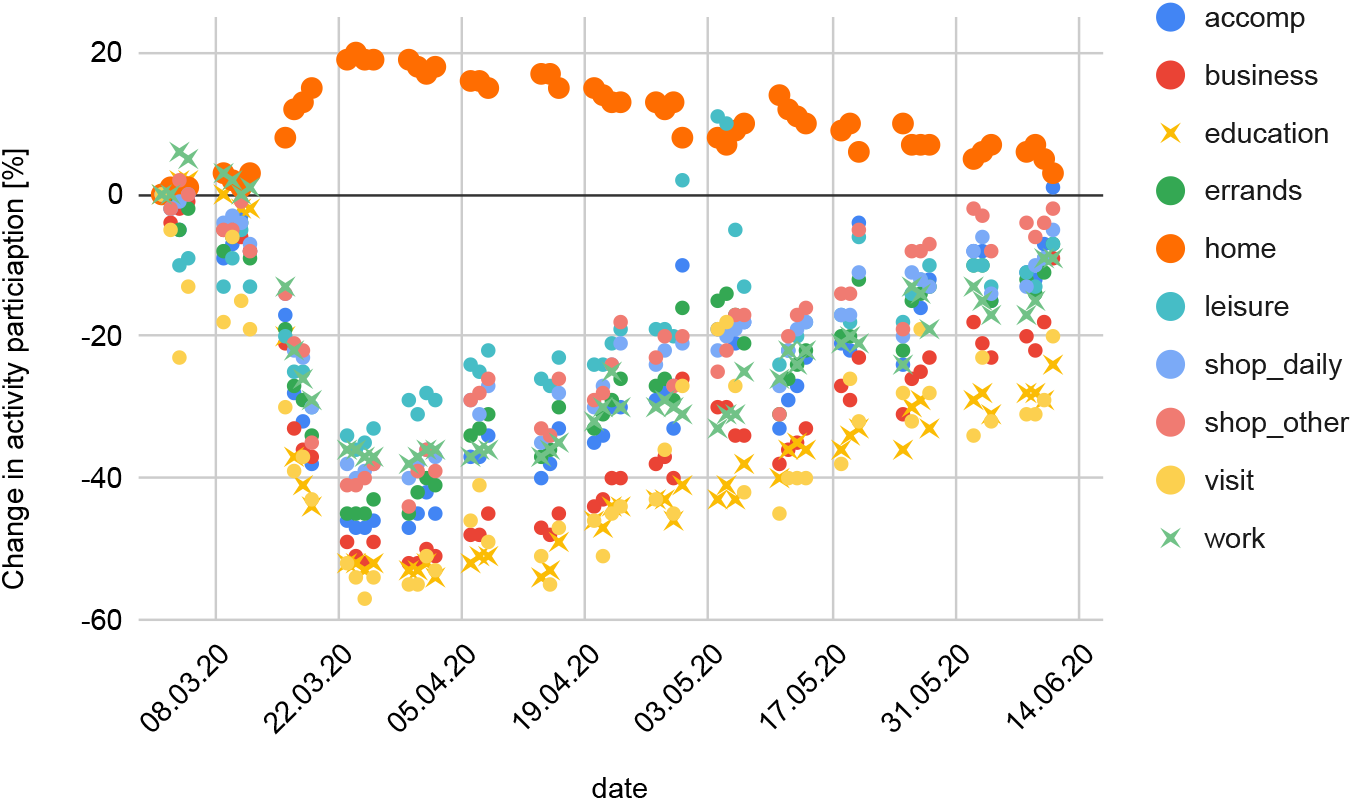
Reduced activity participation over the course of the epidemics in Germany.

**Figure 4.**
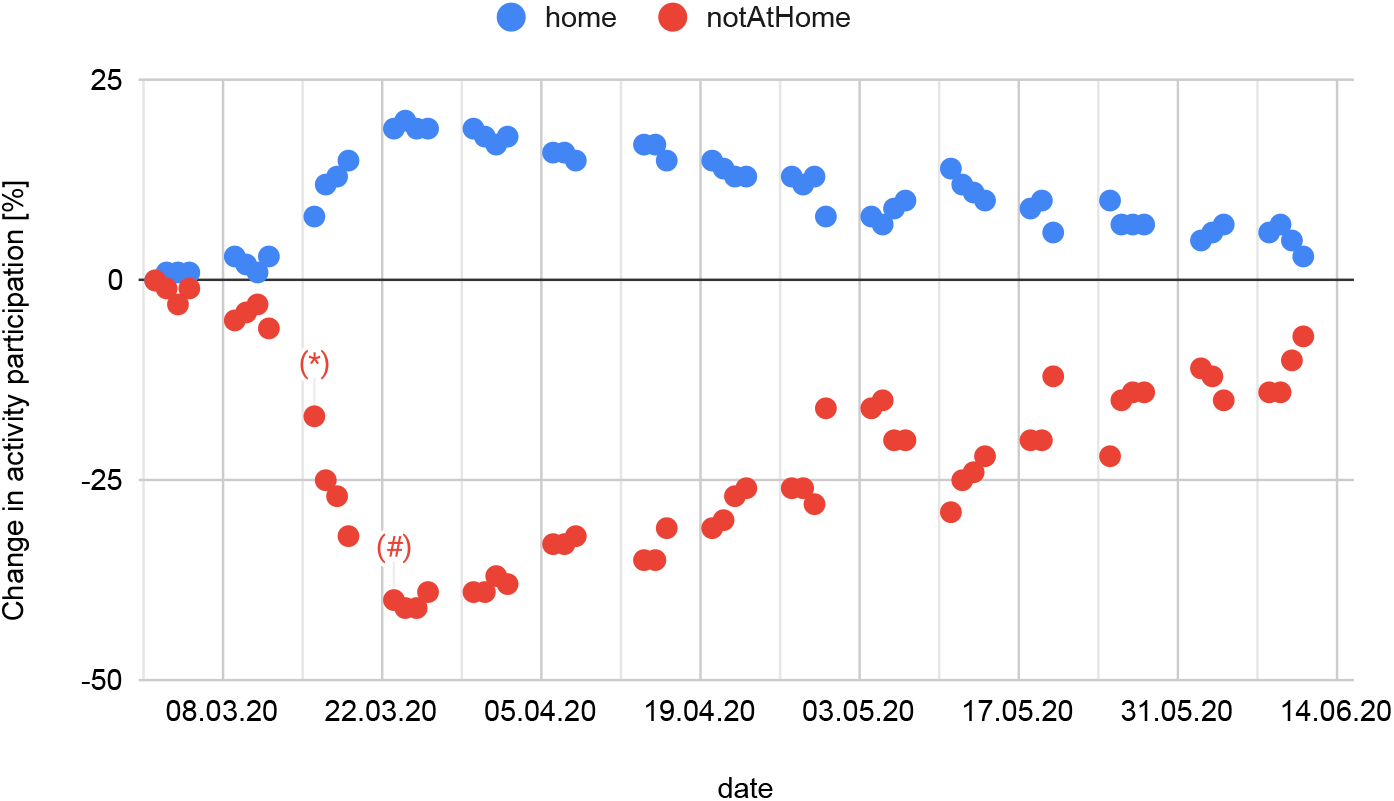
Change in activity participation compared to normal for normal workdays. All out-of-home activities are combined into one number. (*) denotes the first day of closures of schools, clubs, and bars; and (#) the first day of the so-called contact ban which came together with closures of all restaurants and non-essential stores. The data clearly shows that people had reduced their out-of-home activities before the government-ordered closures.

### 2.6 Calibration against case numbers in Berlin

The simulation is calibrated against the Berlin case numbers (Fig. 5, violet dots) [25]. Each reported case comes with two dates: reporting date, and so-called reference date, which refers to the onset of symptoms. Other than Dehning et al. [26], we use the reference date, which is closer to the real dynamics.

**Figure 5.**
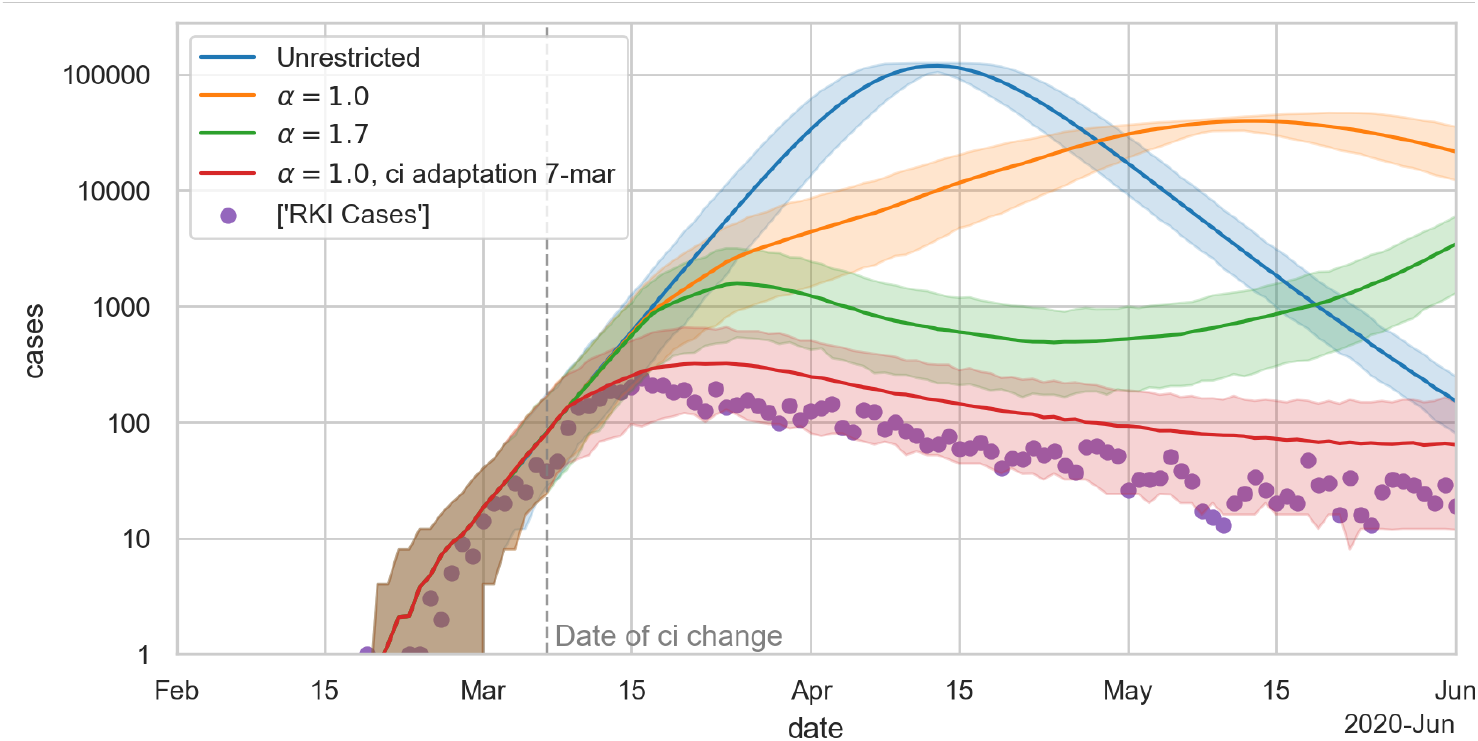
RKI case numbers [25] together with simulated result for different values of a, and final calibration result. The lines show the mean of 300 simulations; the error bands denote 5% and 95% quantiles. The calibration assumes that 1/3 of new infections remain undetected.

Since our simulation explicitly models the state of “showing symptoms”, we can directly compare this to the case numbers (the sampling strategy in Germany at that time was to not test persons without symptoms). Using the model as explained so far leads to the orange curve in Fig. 5. This is clearly too high. Multiplying the reductions in activity participation by a factor (green curve) does not help: This now gets the downward slope right, but the curve implies that the fraction of non-reported cases would be smaller before mid-march than afterwards. Since this is implausible, we set α back to one and introduce a behavioral change point earlier in the simulation, see the red curve. This is consistent with Dehning et al. [26], who report a first change point around Mar/7 which “matches the timing of the first governmental intervention which included cancellations of large events, as well as increased awareness”.

We model this by introducing a change of contact intensity *ci*, early in March, for all out-of-home activities, public transport, and quarantined-at-home. We calibrate the day and magnitude of the *ci* change by minimizing the Root Mean Squared Logarithmic Error (RMSLE) between detected cases and persons showing symptoms in the simulation adjusted by an estimate of 50% unrecorded cases; the two free parameters are the new value of *ci*, and the first day with new value (constrained to be between Mar/3 and Mar/9). As the objective function is expensive to evaluate, we chose the *Tree-structured Parzen Estimator* [27]. The red curve in Fig. 5 shows the final result of the calibration process with the *ci* change point on Mar/7 and *ci* = 0.32. Other than Dehning et al. [26], we do not need a second and third change point; the activity non-participation from our data is sufficient. For a comparison with hospital numbers see Sec. 2.7.

### 2.7 Comparison with hospital numbers in Berlin and resulting statement about undetected cases

Fig. 6 compares our simulation, with *α* = 1 and the additional change point on Mar/7, with reported hospital cases in Berlin [28]. As a part of the calibration process, the hospitalization probabilities of Tab. 1 are multiplied by 1.6. One observes that our model is somewhat “early” when compared to the Berlin hospital admissions. In reaction, one could shift the starting date of the infection process to later. This would improve the timing of the simulated hospitalizations, but the maximum would be lower, which could then be compensated for by a larger multiplier for the hospitalization probabilities. However, then the simulated cases showing symptoms would be lower than the reported case numbers, which seems implausible. Alternatively, one could change the progression model, and assume more time between showing symptoms and hospital admission. This would mean a deviation from the values reported in the literature, which we did not want to do.

**Figure 6.**
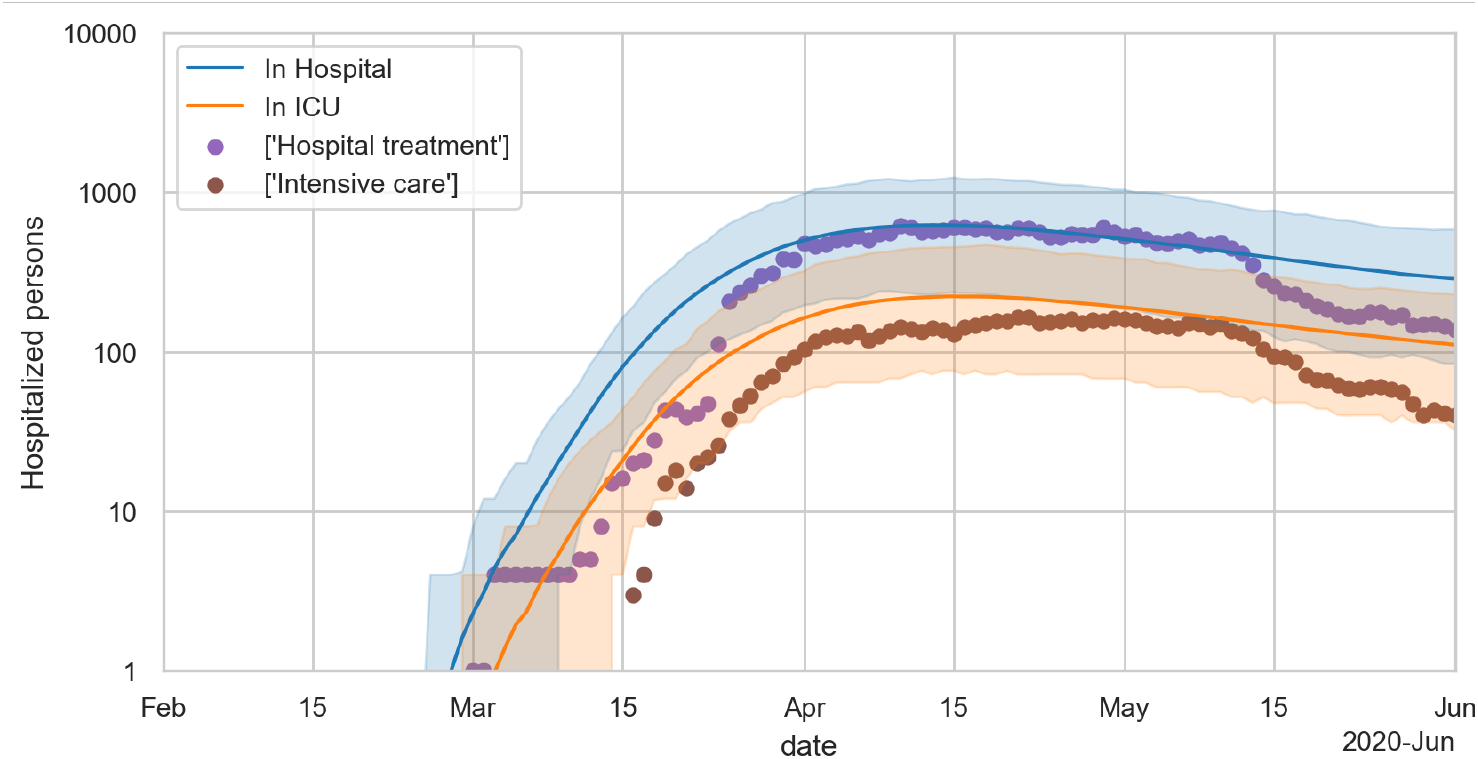
Reported hospital cases [28] together with simulated results. The lines show the mean of 300 simulations; the error bands denote 5% and 95% quantiles.

Given current knowledge, this points to a rather little underreporting in Berlin - since more underreporting would mean an earlier date for the initial infection, which in turn would mean even earlier dates for the hospitalization numbers, contrary to what is observed. This is consistent with other studies [29]. Even with the assumption of little underreporting, however, the disease progression model from the literature is too “fast” when compared to the data in Berlin.

### 2.8 Reductions of the reinfection rate R

In the following, we investigate the contributions of certain non-pharmaceutical interventions towards reducing the reinfection rate *R*. This is obtained by running two different simulations, one without the measure and one with the measure, taking note of the corresponding reinfection numbers, dividing them by each other, and averaging. The following interventions are considered: (1) Certain **types of activities are removed from the infection dynamics** by removing these activity types from each synthetic person’s trajectory, as described above. Note that a 100% reduction of activity participation seems quite unrealistic except for schools; our mobility data (Fig. 4) implies that despite the perceived strength of the measures in Berlin, the drop was never more than 50%. (2) If **activities are removed only with a certain probability**, a random draw is made every time a synthetic person has that activity type in its plan. This means that the model assumes that, say for a 50% work reduction, it will be another 50% subset of persons at work every day. This intervention, in consequence, does not sever infection networks, but just slows down the dynamics. (3) Masks are added into the infection model of Eq.1 by reducing the shedding and/or the intake accordingly: cloth masks reduce shedding and intake to 0.6 and 0.5 of their original values, surgical masks to 0.3 and 0.3, and N95 (FFP) masks to 0.15 and 0.025 [30, 31]. Evidently, it is now possible in our model to assign masks to activity types, to persons, or both. (4) **Contact tracing** is implemented in the following way: If a synthetic person shows symptoms, then after a configurable delay *d* and with a configurable probability *γ*, contact persons of the last 14-days are put into quarantine at home. The parameter *d* parametrizes the delays caused by testing and by tracking down contact persons, the parameter *γ* the probability that a person cannot be contacted or that it does not follow the request to stay at home. Persons in the same household as the symptomatic person are always put into quarantine; contacts during shopping, while using public transport, or with contacts shorter than 15-minutes are ignored.

We determine consequences of interventions by administering them to the unrestricted dynamics (i.e. the blue curve of Fig. 5), on Mar/7. We first collect for each infected person the number of reinfections. These numbers are then averaged over all persons becoming contagious on a certain day; this corresponds to the “case or cohort reproductive number” method of [32]. These numbers are averaged 100 Monte Carlo runs with different random seeds. The resulting *R* values can be plotted over time, see Fig. 7. Evidently, the interventions are visible in these numbers *before* the interventions are implemented, because the reinfections are registered backwards in time to when the person became contagious. However, only from the day of the intervention onward will they show the full effect. To further reduce Monte Carlo noise, we average *ρ* =1 − *R_intervention_/R_base_* over the first 14 days after the introduction of the intervention. For these values, we then compute the *mean* 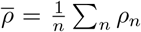, the *standard deviation* 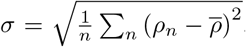, and finally the *standard error* 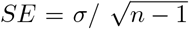 is the number of different simulation runs (Monte Carlo seeds). Evidently, the standard deviation gives information about the variability of the Monte Carlo runs, while the standard error gives information about the probability that the true mean is different from zero.

**Figure 7.**
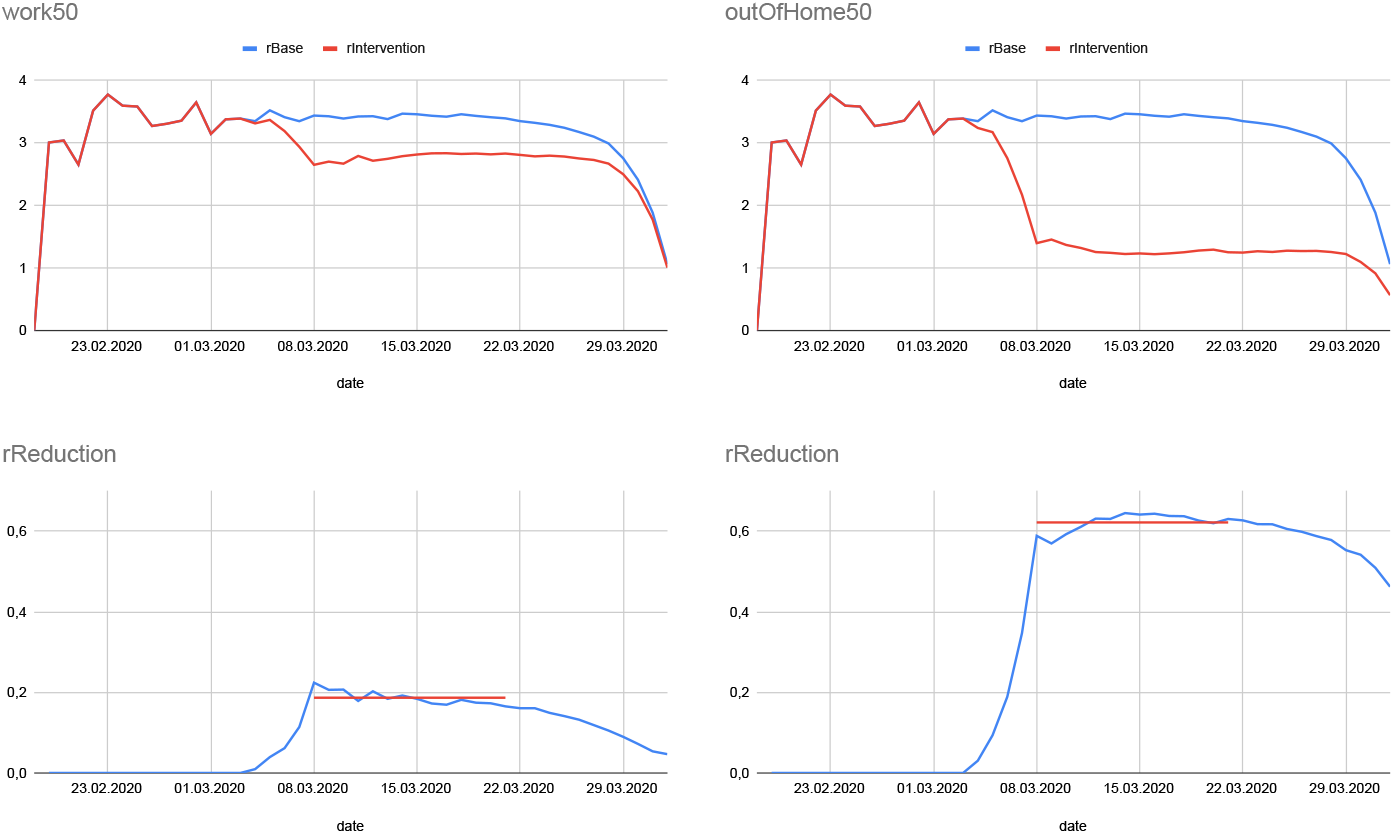
Illustration of determination of different reinfection rates (top) and their ratios (bottom) for two different measures: reduction of out-of-home work activities by 50% (left), and reduction of all out-of-home activities by 50% (right). Each curve is averaged over 100-different Monte Carlo runs. The results given in Table 1 are additionally averaged over the first 14 days after the intervention (red line in bottom figures).

## 3 Results

### 3.1 Behavioral interpretation of government interventions vs mobility data

One striking consequence of the activity participation data (Fig. 4) is that, after the initial government intervention from Mar/7 that cancelled large events and raised awareness, the population reaction in fact *preceded* the government interventions, rather than the other way around. Out-of-home activity participation was already reduced between Mar/7 and Mar/14, *before* the second government intervention that closed schools, clubs, and bars. Similarly, there was a further considerable drop between Mar/14 and Mar/21, again *before* the so-called contact ban (strongly reduced interaction between different households) and closing of all restaurants and non-essential stores in Germany. At least for Berlin and probably for Germany, it is thus not true that the government forced society and the economy to come to a halt; rather, society did this by itself, and the government presumably stabilized or reinforced behavior that was happening anyways.

### 3.2 Reductions of the reinfection rate R

Tab. 2 shows the reductions of the reinfection rate *R* for different policy interventions, as computed by our approach. As simulation output, all of our values are significantly different from zero at the 3a level. One notices the strong reduction of R from reducing all out-of-home activities by 50% - this roughly corresponds to the situation in Germany at the end of March (cf. Fig. 4). We also find that this is a combination of many much smaller contributions: only the reductions of work or of leisure activities reach two digit ranges for the reduction of *R*.

**Table 2.** Additional reduction of R by various non-pharmaceutical interventions.

| Measure | Additional reduction of $R$ by: | Standard deviation | Standard error |
| --- | --- | --- | --- |
| Complete closing of daycare, schools, universities | 6% | 3% | 0.26% |
| Reduction of daycare, school, university attendance by 50% | 5% | 2% | 0.25% |
| Reduction of out-of-home leisure activities by 50% | 15% | 2% | 0.25% |
| Reduction of out-of-home shopping activities by 50% | 3% | 3% | 0.26% |
| Reduction of out-of-home work activities by 50% | 19% | 3% | 0.31% |
| Reduction of all out-of-home activities by 50% | 62% | 2% | 0.18% |
| Cloth masks are worn by 50% of persons and surgical masks by an additional 10% while shopping and in public transport | 4% | 2% | 0.25% |
| N95 masks are worn by 90% of persons while shopping and in public transport | 7% | 3% | 0.28% |
| N95 masks are worn by 90% of persons at all out-of-home work activities | 20% | 3% | 0.26% |
| Contact tracing (with $d = 2$ and $\gamma = 0.5$) followed by quarantine-at-home | 33% | 3% | 0.28% |
| Contact tracing (with $d = 2$ and $\gamma = 0.75$) followed by quarantine-at-home | 40% | 3% | 0.33% |

Similarly, masks help, but a two digit impact can only be reached when wearing masks at work (where our measure is maybe better translated as “90% of people wear N95 masks at work *or use single person offices*”).

Also contact tracing followed by quarantine-at-home has a strong impact. Evidently, this assumes that the contact tracing system is not overwhelmed by large infection numbers. We also find that even a system with infinite tracing capacity results in a finite reduction of R. In consequence, if contact tracing is not sufficient, it needs to be supported by other measures.

Assuming that the initial changes in behavior from Mar/7 are changes that will remain stable over the epidemics and the remaining *R* is around 2, then still a combination of relatively many measures is needed to bring *R* to below one. Note that this needs to be done multiplicatively, i.e. 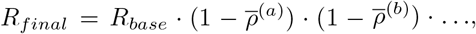 where (*a*), (*b*),... are the different measures. If contact tracing is part of this package, then if the contact tracing system becomes overwhelmed, situation where R was maybe slightly above one will suddenly jump to one where *R* is about 30% to 40% larger. The result will be a strong acceleration of the exponential growth dynamics, and fairly strong measures will have to be taken to get *R* back to below one. We think that successful contact tracing is a plausible explanation why the situation is currently (middle of July) under control in Germany despite the relatively open regime compared to other countries.

## 4 Discussion

Arguably, compartmental models are the mainstay of epidemiological modelling. Our approach, in contrast, follows individual synthetic persons. These individual persons can be enriched by person-centric attributes such as age or individual risk factors. Disease progression is individual, taking into account these demographic and other person-centric attributes. Similar to compartmental models, the base reinfection rate and the starting date need to be calibrated from case numbers. However, both the spatial and the social interactions in our model come directly from data. Also, behavioral reductions in activity participation come directly from data. Mechanical aspects such as the wearing of masks by certain persons and/or at certain activity types can be integrated very simply into the model, by reducing virus shedding, virus intake, or both. Travel in public transport is already integrated. Organizational suppression approaches, such as contact tracing, can be simulated mechanically, thus extracting information about the allowed delays between symptom onset and reaching contacts, the failure rate, etc.

We were able to bring this up quickly: Coding of the infection code was started at the end of Feb/2020; our first preprint is from 20/Mar/2020 [33]; our first report to the government is from 8/Apr/2020 [34]; we have reported to the government regularly since then^[1]^. Evidently, we were drawing from our experience and expertise with person-centric travel models. Still, it means that given the right experience and data availability, the method is not overly heavyweight, and then has many advantages over compartmental models.

The basic behavior of the model is like that of any S(E)IR model, i.e. exponential growth until a sufficient share of the population is immune, followed by exponential decline (cf. blue line in Fig. 5). Also the beginning and the speed of the growth are calibrated in similar ways. In most models, however, interventions such as reductions in out-of-home activity participation, masks, or contact tracing, need to be parametrized into parameter changes of the S(E)IR model, most notably the infection rate [26, 35, 36, 37]. The only models that use human activity patterns directly that we are aware of are the three models described in [15]. Out of these, we are aware of an application to COVID only by the Imperial College model [16]. Their results are roughly in line with ours. That model, at the time, used a doubling of cases every 10 days; reality, with a doubling every 3 days, was possibly even more dramatic than their predictions. However, their model was purely predictive, i.e. they did not use mobility data to gauge the actual reductions in activity participation.

Tab. 3 is an attempt to extract “additional reductions to *R*” from other studies. One immediately finds two issues: (A) The categories are not well aligned. For example, “small gathering cancellation” refers to gatherings with 50 persons or less, while other studies cancel gatherings *larger* than a certain number. Again other studies just consider a “gathering ban”, but at the same time have “event ban” and “venue closure” as separate items. (B) Even where the categories are well aligned, the resulting numbers vary widely: “schools closed” goes from 5% to 50%, “national lockdown” goes from 0 to 81%.

**Table 3.** Percent reduction of R in other studies. Rounded to integers.

| Measure | [38] | [39]<br>“CC” | [39]<br>“LASSO” | [40] | [41] |
| --- | --- | --- | --- | --- | --- |
| Schools closed | 50 | 16 | 34 | 8 | 5 |
| Some businesses suspended | 26 |  |  |  |  |
| Most businesses suspended | 34 |  |  |  |  |
| Work ban |  |  |  | 31 |  |
| Gatherings limited to $\leq 1000$ | 16 | | | | |
| Gatherings limited to $\leq 100$ | 17 | | | | |
| Gatherings limited to $\leq 10$ | 28 | | | | |
| Mass gathering cancellation ($> 50$) | | 27 | 0 | | |
| Small gathering cancellation ($\leq 50$) | | 17 | 23 | | |
| Event ban |  |  |  | 23 | 5 |
| Gathering ban |  |  |  | 34 |  |
| Venue closure |  |  |  | 36 |  |
| Stay-at-home order with exemptions | 14 |  |  |  |  |
| National lockdown |  | 0 | 8 | 5 | 81 |
| Mask-wearing | 0* |  |  |  |  |
| Increase availability of person protective equipment |  | 9 | 12 |  |  |

\* not significantly different from zero

In part, this is a consequence of the fact that the interventions are not standardized: For example, the number of exemptions in what is called a lockdown varied quite a lot between countries. Additionally, the transmission mechanisms vary, so even if the concept may be the same, the execution and thus the effect may be quite different between countries.

For example, our reductions to *R* caused by school closurs come out much lower than most other values, in particular those of Brauner et al. [38]. We attribute this to the following two elements: First, the model by Brauner et al. has no additional change point on Mar/7. In consequence, their approach has to assign all changes in the infection dynamics to the school closures. The school closures in Berlin were, however, one week later, with Mar/12 (fri) or Mar/15 (mon) as the last day of school; too late to explain the first change in the infection numbers. Also, Dehning et al. [26] have an additional change point on Mar/7, corroborating its existence in Germany. Second, other than both Brauner et al. and Dehning et al., we have the mobility data of Fig. 4 at our disposal. It is clear that there was considerably more societal adaptation around the weekend of Mar/13-14 than just keeping children at home. Brauner et al. themselves write that “the closure of schools … may have caused … behaviour changes. We do not distinguish this indirect signalling effect from the direct effect”. Additionally, in Germany, children staying at home will force their parents to stay at home, thus forcing them into home office. In consequence, some of this may not be signalling, but causal secondary effects. In consequence, our model is more differentiated: What Brauner et al. attribute to the school closures alone is in our model attributed to a combination of school closures, behavioral changes, and the reduction of various other out-of-home activities. Thus, all of the values may be correct: The pure effect of school closures in western countries (with relatively few young people) may not be larger than 5%, but the measurable consequence for *R* when governments closed schools as their first intervention presumably indeed was much larger.

We have checked our relatively large reductions of R for masks multiple times. They are a consequence of the assumption that N95 masks reduce intake to 2.5%, taken from [30]. The review article [31] comes up with about 5%, a factor of two larger, but still displaying a very large reduction. The same paper [31] also shows that “masks” without a specification of the type has much less of an effect. Finally, there may be the issue that lay people may not be able to use N95 masks at full efficiency. In consequence, our results have to be interpreted once more “mechanically”: They are plausible under the assumption that the fraction of people specified in the model is indeed able to use N95 masks effectively.

Clearly, data-driven mechanical models such as ours help clarifying the categories since we can exactly specify what we mean by closing some activity type or wearing a mask at certain activity types. Also, we can differentiate between the transmission from political decision to behavioral execution vs. the consequences of the behavioral execution to the infection dynamics. Finally, we can mechanically include organizational approaches such as contact tracing.

An issue that needs to be investigated further is our setting of the contact intensity *ci* uniformly to one. It is quite clear that *ci* cannot be simply connected to the activity type; in particular, the person density and thus the contact intensity in public transport is much higher during peak hours than outside peak hours. Possibly, the contact intensity can be related to room sizes and air exchange rates [42]; this will be a subject of future work.

## 5 Conclusions

We combine a person-centric human mobility model with a mechanical model of infection and a person-centric disease progression model into an epidemiological simulation model. Different from other models, we take the movements of the persons, including the intervening activities where they can interact with other people, directly from data. For privacy reasons, we rely on a process that takes the original mobile phone data, extracts statistical properties, and then synthesizes movement trajectories from the statistical properties; one could use the original mobile phone trajectories directly if they were available. The model is used to replay the epidemics in Berlin. This allows important insights into the societal transmission from government actions to mobility behavior to infection dynamics. Importantly, it turns out that the population started reducing its out-of-home activities *before* the government asked/ordered the population to do so. The model is then used to evaluate different intervention strategies, such as closing educational facilities, reducing other out-of-home activities, wearing masks, or contact tracing, and to determine differentiated percentage changes of the reinfection number R per intervention. An important insight is that successful contact tracing followed by quarantine-at-home results in a rather large reduction of the reinfection rate R. In consequence, if contact tracing becomes overrun, rather drastic lockdown measures are necessary to bring R back to below one.

## Data Availability

Access to computer code and output data is provided in the paper. Access to input data is unfortunately on "reasonable" request only, as specified in the paper.

https://github.com/matsim-org/matsim-episim

https://svn.vsp.tu-berlin.de/repos/public-svn/matsim/scenarios/countries/de/episim/output-data/stability1.zip

https://svn.vsp.tu-berlin.de/repos/public-svn/matsim/scenarios/countries/de/episim/output-data/stability2.zip

https://svn.vsp.tu-berlin.de/repos/public-svn/matsim/scenarios/countries/de/episim/output-data/interventions0703/

https://covid-sim.info/

## Ethics approval and consent to participate

Not applicable

## Consent for publication

Not applicable

## Availability of data and materials

For computer code see https://github.com/matsim-org/matsim-episim. Figs. 5 and 6 were computed with version ab6515b557c293af24eda254a79b5d4d7ccb4bd9 of the code, started with command

`java -jar matsim-episim-1.0-SNAPSHOT.jar runParallel \`

`--setup org.matsim.run.batch.StabilityRuns \`

`--params org.matsim.run.batch.StabilityRuns$Params`

The output data used for the figures can be retrieved at https://svn.vsp.tu-berlin.de/repos/public-svn/matsim/scenarios/countries/de/episim/output-data/stability1.zip and https://svn.vsp.tu-berlin.de/repos/public-svn/matsim/scenarios/countries/de/episim/output-data/stability2.zip.

Fig. 7 and Tab. 2 were computed with version dad3a44989c2ab2131c17fc1a6b533906012d47f of the code, started with command (number of seeds were set to 100 in Params class)

`java -jar matsim-episim-1.0-SNAPSHOT.jar runParallel \`

`--setup org.matsim.run.batch.Interventions \`

`--params org.matsim.run.batch.Interventions$Params`

Post processing was done with

`java -jar matsim-episim-1.0-SNAPSHOT.jar org.matsim.analysis.SMRValuesFromEvents`

The output data used for Fig. 7 and Tab. 2 can be retrieved at https://svn.vsp.tu-berlin.de/repos/public-svn/matsim/scenarios/countries/de/episim/output-data/interventions0703/.

The input data to the simulations (synthetic mobility traces) are available from Senozon but restrictions apply to the availability of these data, which were used under license for the current study, and so are unfortunately not publicly available. Data are however available from the authors upon reasonable request and with permission of Senozon. Also, the code in the repository is runnable with a synthetic version of such data.

The only input that is absolutely necessary to run the code are the initial activity-based person trajectories, which can, e.g., be obtained from mobile phone data, or from so-called activity-based models in transport.

More results and visualizations are available under https://covid-sim.info.

## Competing interests

MB and KN own shares of Senozon. Beyond that, the authors declare that they have no competing interests.

## Author’s contributions

Conceptualization KN; data curation AN, MB, CR, SAM, RE, TS, BC; funding acquisition KN; investigation SAM; methodology SAM, KN, CR; project administration RE; software CR, SAM; visualization BC, SAM, CR, RE, KN; writing KN, SAM, CR.

## Acknowledgements

We thank Kai Martins-Turner and Dominik Ziemke for discussions, and Timo Smieszek for comments on an early draft. The work on the paper was funded by the Ministry of research and education (BMBF) Germany (01KX2022A) and TU Berlin; regular reports can be found through this search:

https://depositonce.tu-berlin.de/simple-search?query=modus-covid. Zuse Institute Berlin (ZIB) provided CPU time.

## Footnotes

[1] Cf. https://depositonce.tu-berlin.de/simple-search?query=modus-covid

## Notes

### Competing Interest Statement

KN and MB own shares of Senozon, the company which generates the synthetic movement traces.

### Summary of Updates

some material moved from conclusion to discussion; discussion expanded; stylistic changes

